# A systematic narrative review of coroners’ prevention of future deaths reports (PFDs): a potential metric for patient safety in hospitals

**DOI:** 10.1101/2023.05.19.23290187

**Authors:** Benjamin T. Bremner, Carl Heneghan, Jeffrey K. Aronson, Georgia C. Richards

## Abstract

Patient harm due to unsafe healthcare is widespread, potentially devastating, and often preventable. Hoping to eliminate avoidable harms, the World Health Organization (WHO) published the Global Patient Safety Action Plan in July 2021. The UK’s National Health Service relies on several measures, including “never events”, “serious incidents”, patient safety events, and coroners’ prevention of future death reports (PFDs) to monitor healthcare quality and safety. We conducted a systematic narrative review of PubMed and medRxiv on 19 February 2023 to explore the strengths and limitations of coroners’ PFDs and whether they could be a safety metric to help meet the WHO’s Global Patient Safety Action Plan. We identified 17 studies that investigated a range of PFDs, including preventable deaths involving medicines and an assessment during the COVID-19 pandemic. We found that PFDs offered important information that could support hospitals to improve patient safety and prevent deaths. However, inconsistent reporting, low response rates to PFDs, and difficulty in accessing, analysing, and monitoring PFDs limited their use and adoption as a patient safety metric for hospitals. To fulfil the potential of PFDs, a national system is required that develops guidelines, sanctions failed responses, and embeds technology to encourage the prevention of future deaths.

## Introduction

Patient harm due to unsafe care is a growing global public health challenge that demands an urgent international response. In July 2021, the World Health Organization (WHO) published the Global Patient Safety Action Plan, with the intention of eliminating avoidable harms in healthcare^1^. In the UK, the National Health Service (NHS) has several initiatives and measures in place to monitor and improve patient safety, including “never events”, “serious incidents”, coroners’ prevention of future death reports (PFDs), and patient safety events reported by patients, the public, and staff^2^. However, it has been estimated that over 10,000 adult deaths in English hospitals are preventable each year^3^.

Death is the most severe and objective marker of harm, making it the most used primary outcome in research and healthcare settings worldwide. However, not all deaths are inevitable. Treatment and prevention are two mechanisms by which avoidable deaths can be averted. According to the Office for National Statistics (ONS), *preventable mortality* describes a death that can be avoided ‘before onset of disease or injury… through effective public health and primary interventions’^4^, and *treatable mortality* is death that can be avoided ‘after onset of a disease… through timely and effective healthcare interventions’ to reduce case-fatality^4^.

### Patient safety metrics

Each year in the National Health Service (NHS), 32,000 records of so-called patient safety events (i.e. harms) are reviewed^2^. The NHS estimates that 160 lives and £13.5 million in treatment costs are saved every year because of the incident review and response efforts of the NHS Patient Safety Strategy^5^. Records are provided from established sources, including Serious Incident reports, Never Events, events that are recorded on the National Reporting and Learning System (NRLS), and coroners’ Prevention of Future Deaths reports (PFDs)^2^. NHS England and affiliated organisations publish summaries and reports of Never Events at regular intervals^6,7^. Furthermore, patient safety incident reports are stored in the NRLS database, with organisational and national summaries published by NHS England and NHS Improvement^8^. A systematic search of the literature showed that most published research on harms refers to ‘never events’ (Figure 1, Table S1), and PFDs were an underexploited resource. To explore the potential of PFDs, we conducted a systematic narrative review of studies that investigated PFDs.

**Figure 1:**
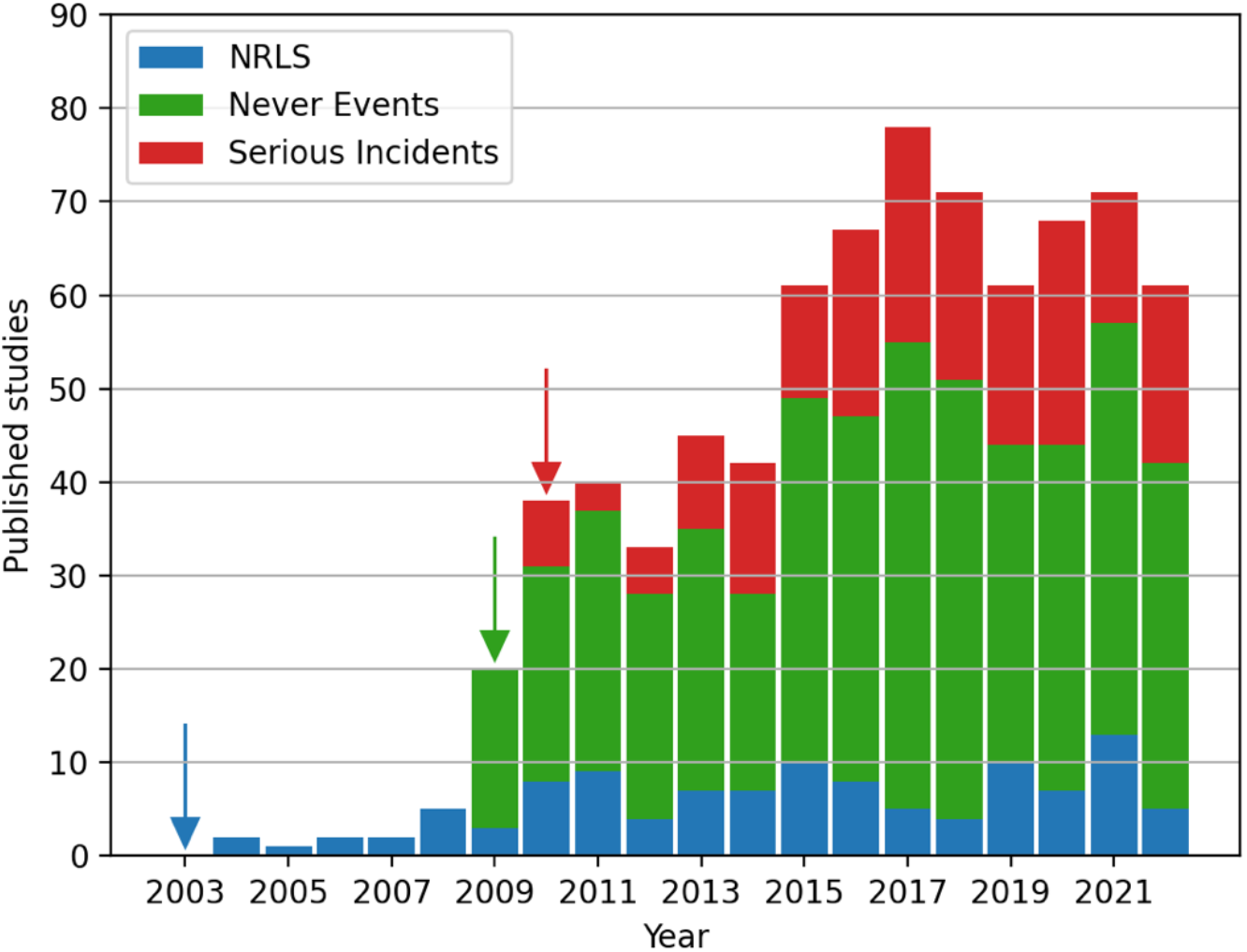
Bibliometric analysis of patient safety events (i.e. harms) as indexed in PubMed from their dates of introduction (arrows) to 16 November 2022, including the National Reporting and Learning System (NRLS; blue), which was introduced in 2003 (blue arrow), Never Events (green), which were introduced in 2009 (green arrow), and Serious Incident reports (red), which were introduced in 2010 (red arrow)^12^. The search strategy is summarised in Supplement Table S1.

### What are PFDs?

Coroners issue Prevention of Future Death reports (PFDs) when they consider it appropriate to share information detailing actions to be taken to prevent further deaths. PFDs are mandated under The Coroners and Justice Act 2009^9^ and The Coroners (Investigation) Regulations 2013^10^. Three processes are involved in issuing PFDs (Figure 2). Once a PFD is written, it is uploaded to the Courts and Tribunal Judiciary website^11^, and assigned to a “*category*” or multiple categories of deaths.

**Figure 2:**
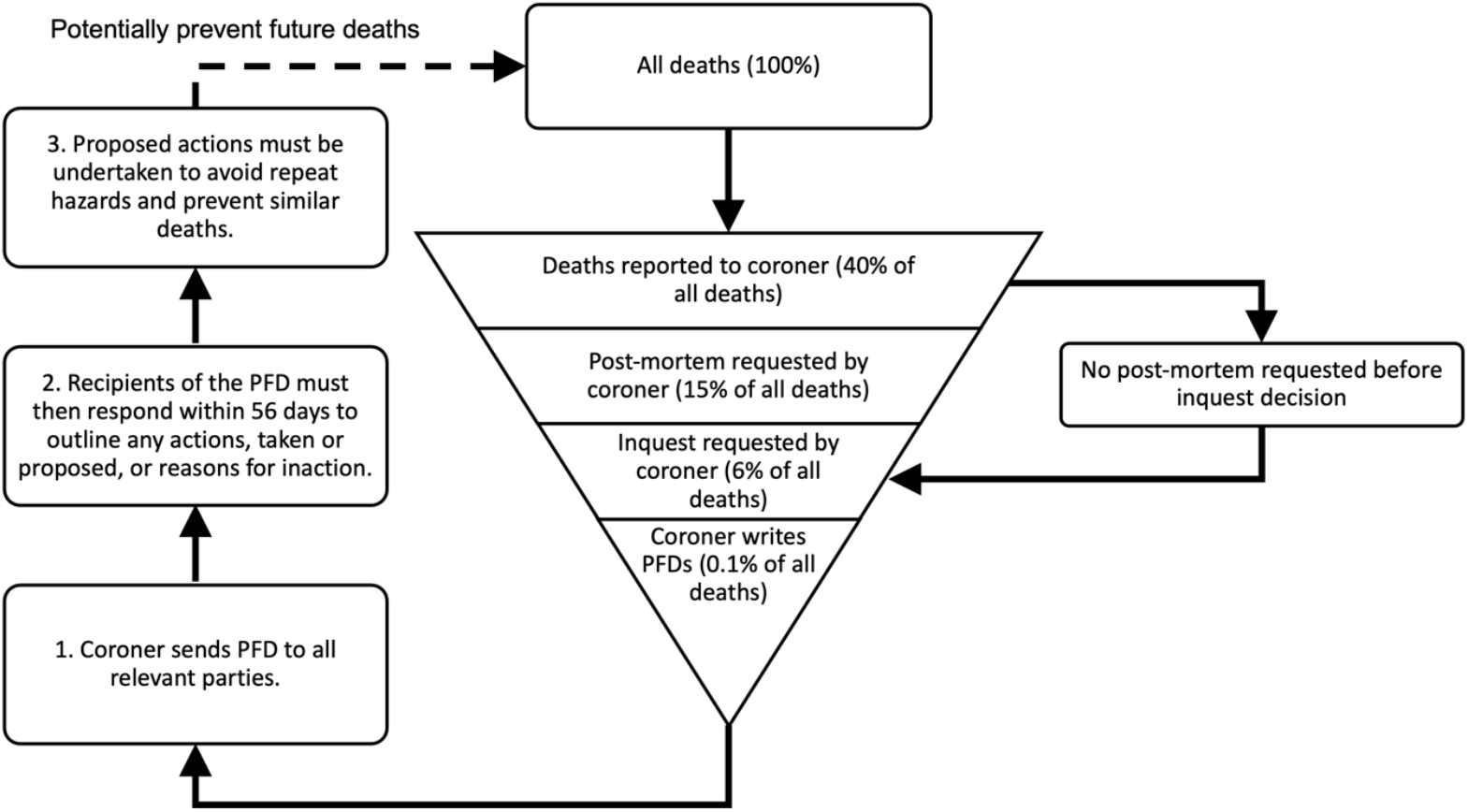
The Prevention of Future Deaths reports (PFD) process in the context of all deaths in England and Wales, adapted from Zhang & Richards 2022^13^. When an unnatural or unclear death occurs, it may be reported to a coroner. Further investigations, including a post-mortem and inquest, may be requested by the coroner to explore the circumstances and causes of death. After the inquest, if the coroner deems that the death could have been prevented, it is their duty to write a PFD and send it to individuals and/or organisations capable of actions^10^. Recipients of PFDs must respond within 56 days under the statute^10^.PFDs and their responses may be published on the Courts and Tribunal Judiciary website^11^. The percentage of deaths proceeding through each stage was reported in the Chief Coroner’s Annual report for 2019^14^.

## Methods

We conducted a systematic narrative review to identify literature relating to PFDs. We searched PubMed and medRxiv on 19 February 2023 and conducted forward (citations) and backward (reference lists) screening of included studies. Search terms and the search strategy are available in Supplement Table S2 and S3. For medRxiv, the first two pages of the database were screened to identify unpublished literature.

Studies were screened independently by one reviewer (BTB), cross-checked by a second reviewer (GCR), and shared with a clinical pharmacologist who is an expert in the field (JKA). Studies were included if they reported the use of PFDs as a source of data and/or discussed the strengths and limitations of PFDs in practice and were published in English and related to the English and Welsh coronial and healthcare system regardless of study design. Research letters, commentaries, and editorials were excluded.

Data were extracted and synthesised by one reviewer (BTB) and cross-checked by a second reviewer (GCR). Data relating to the types of PFDs, reported outcomes, and strengths and limitations of included studies were summarised narratively. No meta-analysis and/or quality assessment was performed. Graphs were created with matplotlib.pyplot and numpy packages in Python using Spyder, and flow diagrams were created using Microsoft PowerPoint.

This review was not preregistered, and no review protocol was prepared, owing to the narrative nature and study design. The findings were reported according to the Preferred Reporting Items for Systematic Reviews and Meta-Analyses (PRISMA) checklist where applicable.

## Results

After screening and removing duplicates, 17 studies^13,15–30^ that examined PFDs in England and Wales were included (Figure 3).

**Figure 3:**
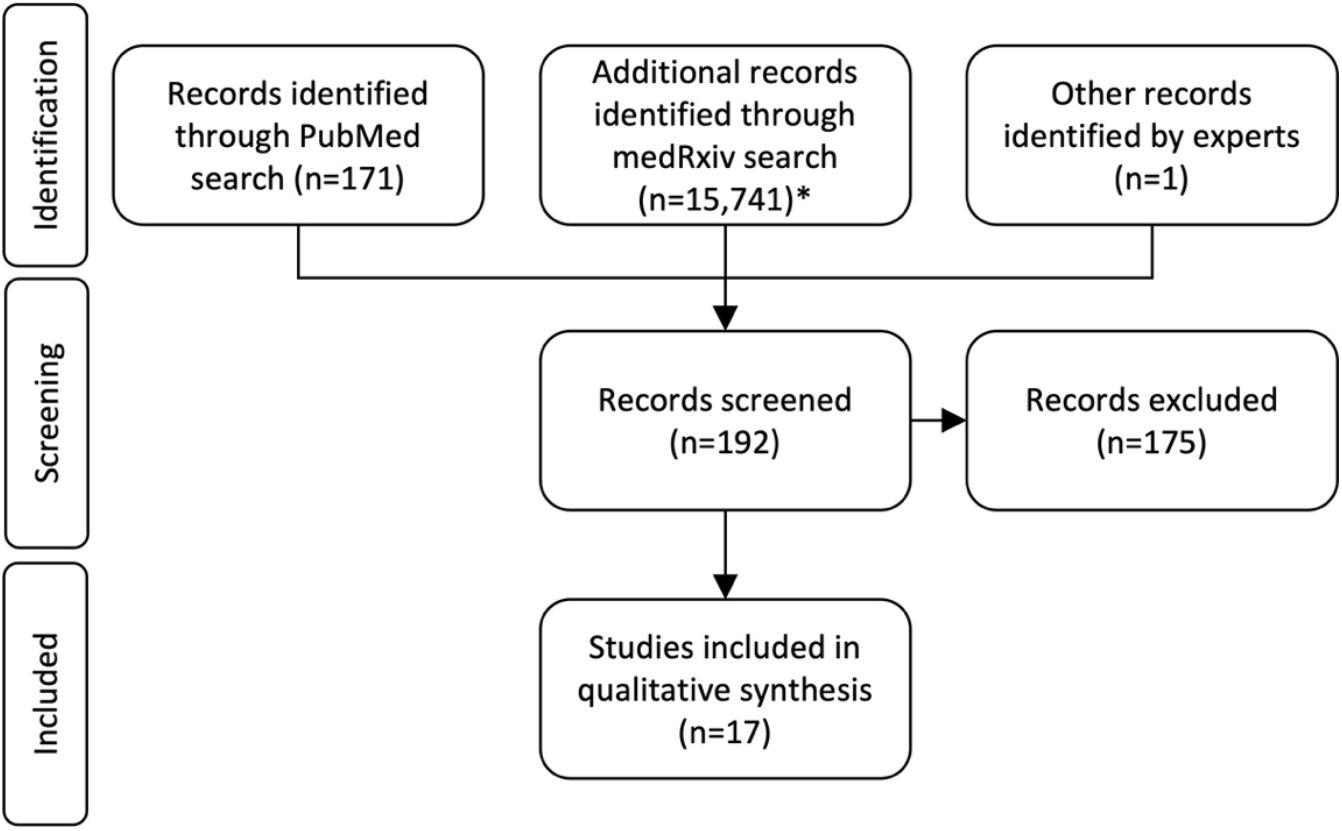
Flowchart of the search conducted to identify studies analysing coroners’ Prevention of Future Deaths reports (PFDs). The search strategy used for PubMed and medRxiv databases are available in Supplement Table S2 and Table S3. *For medRxiv, the first two pages (20 results) were screened to identify unpublished literature.

Eight studies (47%) explored PFDs describing medicines and drugs that had caused or contributed to deaths^15,16,19,24–26,29,30^, two studies assessed PFDs involving chemical products, including paraffin-based emollient creams and alcohol-based hand sanitisers^21,28^, one study examined PFDs involving diagnostic errors^27^, four publications explored healthcare-related PFDs^17,18,20,22^, and two studies analysed PFDs using a selection of categories from the Courts and Tribunals Judiciary website^13,23^ (Table 1). Across these studies, preventable deaths often occurred in hospitals. An analysis of all available PFDs between July 2013 and June 2022 (n=4001) showed that hospital-related deaths were the most common category (44%; n=1772 PFDs) reported on the Courts and Tribunals Judiciary website^13^. We therefore focused on the strengths and limitations of PFDs as a metric for assessing patient safety in hospitals.

**Table 1:** Summary of included studies investigating coroners’ Prevention of Future Deaths reports (PFDs) indexed in PubMed and medRxiv from inception until 19 February 2023, ordered alphabetically by category.

| Study ID | Population/types of PFDs | Outcomes |
| --- | --- | --- |
| **Medicine and drug-related** |  |  |
| Anis et al. 2022<sup>29</sup> | 113 CVD-related PFDs involving anticoagulants between Jul 2013 and Dec 2019 | Warfarin, enoxaparin, and rivaroxaban were most involved in CVD-related PFDs. Poor systems, communication and failures in accurate medical records were the most common concerns reported by coroners. PFDs were most often sent to NHS trusts, hospitals, and general practices, but 60% had not received responses. |
| Aronson et al. 2022<sup>30</sup> | 17 PFDs where medicines or non-medicinal chemicals were purchased online and contributed to death, dated between Nov 2018 and Dec 2019 | In two PFDs, products were obtained from the dark web. Mental health and drug dependence/abuse were the most common contributory factors. Prescription-only medications were the most involved drugs. Concerns included ease of obtaining medicines, regulation of supply and importation of medicines, including the regulation of the dark web, and lack of limits on the amount and frequency of orders. 19 of 21 (90%) recipients did not respond to the coroner within the legal time limit. |
| Cox & Ferner 2021<sup>26</sup> | Two PFDs associated with tramadol | PFDs highlighted the dangers of repeat prescribing of tramadol, the importance of communicating its risks, and the need for evidence-based solutions to prevent future harms. |
| Dernie et al. 2022<sup>16</sup> | 219 PFDs involving opioids | For every opioid-related death, a median of 33 years of life were lost. Morphine, methadone, and heroin were most often involved. Coroners' concerns were frequently related to systems and protocols and maintaining accurate medical records and plans for patients taking opioids. Concerns were most often addressed to NHS organisations. Responses to PFDs were poor (47% overall) and their lessons were only disseminated locally. Wide geographical variation in the writing of reports was noted. |
| Ferner et al. 2019<sup>25</sup> | Responses to 69 of 99 PFDs relating to medicines, published between Apr 2015 and Sep 2016 from 106 organisations | Common actions taken by organisations were staff education or training and changes to processes or policy, although some organisations felt existing policies were sufficient. For 53 reports with relevant dates available, the median time for a coroner to issue a report was 240 days after the date of death. The median time taken for organisations to respond to PFDs was 53 days. |
| Ferner et al. 2018<sup>24</sup> | 99 PFDs in which medicines or the medication process was identified, published between Apr 2015 and Sep 2016 | Anticoagulants, antidepressants, and drugs of abuse were the most mentioned medicines. Adverse reactions to prescribed medications, omission of necessary treatment, failure to monitor treatment, and poor systems were the most highlighted concerns. Concerns were related to issues in education or training, lack of clear guidelines or protocols, and failure to implement existing guidelines. |
| France et al. 2022<sup>15</sup> | 704 PFDs involving medicines, published between Jul 2013 and Feb 2022 | 1 in 5 PFDs involved medicines. Of the 716 deaths involving medicines, it was estimated that 19,740 years of life were lost. Opioids, antidepressants, and hypnotics were the most common medicines involved. Concerns raised by coroners were most often related to patient safety and communication. The most common recipients of medicines-related PFDs were NHS England, the DHSC, and the MHRA. 51% of expected responses were not available. |
| Thomas & Richards 2021<sup>19</sup> | One PFD attributed to an NSAID, diclofenac | Concerns raised by coroners, such as missed opportunities for further testing and observations, and systemic failures in documentation and transmission of information. |
| **Chemical product-related** |  |  |
| Bilip & Richards 2021<sup>28</sup> | One PFD relating to paraffin-based emollient creams | Coroners' concerns were that there had been frequent fires when paraffin-based creams were in use, involving mostly elderly residents; the wide availability of paraffin-based creams and lack of awareness of their fire risk; the risk of potential build up on clothing and the lethal outcomes of these fires. |
| Richards 2021<sup>21</sup> | Two PFDs describing deaths from ingesting alcohol-based hand sanitisers | Concerns raised by coroners were identified and used to form eight recommendations to prevent intentional and accidental ingestion of alcohol-based hand sanitisers in healthcare and community settings. |
| **Diagnosis-related** |  |  |
| Cooper et al. 2021<sup>27</sup> | Nine PFDs involving diagnostic errors related to GP services in or alongside emergency departments, published between 2013 and 2018 | Potential priority areas for improvement were considered, including difficulty in identifying appropriate patients for GP services; under-investigation and misinterpretation of diagnostic tests; inadequate communication and referral pathways. |
| **Healthcare-related** |  |  |
| King & Benbow 2022<sup>22</sup> | 159 PFDs in two categories, hospital-related and community health care and emergency service-related deaths, published before May 2021 | Analysis showed significant variability in the detail and quality of reporting and in the frequency of reporting between different geographical regions. No responses were published for almost 20% of reports. |
| Leary et al. 2021<sup>18</sup> | 710 healthcare-related PFDs, published between 2016 and 2019 | 36 PFDs highlighted that they were sending PFDs to the same organisation regarding the same or similar concerns. The primary themes were deficits in skill or knowledge, missed, delayed, or uncoordinated care, communication and cultural problems, systems issues, and lack of resources. |
| Swift et al. 2022<sup>20</sup> | 23 PFDs relating to SARS-CoV-2 published between Jan 2020 and Jun 2021 | The most common coroners' concerns were communication problems and failure to follow protocols. NHS organisations and the Government were the most common recipients of PFDs; however, response rates were poor. There was significant geographical variation in the reporting of PFDs. |
| Van Dellen et al. 2022<sup>17</sup> | The role of psychiatrists in the coronial process | Thoughtful responses from psychiatrists who are subjects of criticism towards questions about their learning from an incident can avoid the creation of PFDs. Furthermore, a psychiatrist can explain why a GMC referral is unnecessary in a PFD. Submissions can refer to evidence and nominate individuals or bodies to whom the report should be addressed. |
| **Mixed categories\*** |  |  |
| Fox & Jacobson 2021<sup>23</sup> | 50 'recently' published PFDs as of 10 Jun 2020 for categories including child deaths, alcohol, drugs and medications, and railways | Coroners generated PFDs at significantly different rates. Only 37% of PFD addressees responded. Larger organisations were more likely to respond than smaller ones. Robust actions appeared in even fewer responses. A sex imbalance, with a female minority, existed in the PFDs, including neonates. |
| Zhang & Richards 2022<sup>13</sup> | All available PFDs, published between Jul 2013 and Jun 2022 (n=4001) | The most common category of PFD was 'hospital (clinical procedures and medical management)' related deaths. However, 73% of deaths on the Courts and Tribunals Judiciary website were not categorised. There was significant geographical variation in PFD writing: 20 coroners were responsible for 30% of all PFDs. Over 450 coroners had issues with naming. Problems with date formats and coroners' areas were also prevalent. 36% of PFDs did not have an available response. |

PFD: prevention of future death report; CVD: cardiovascular disease; NHS: National Health Service; MHRA: Medicine and Healthcare products Regulatory Agency; DHSC: Department of Health and Social Care. \*Categories are determined by the Chief Coroner's Office when shared on the Courts and Tribunals Judiciary website

### Strengths of PFDs

Studies investigating PFDs have highlighted that they contain rich information and provide essential lessons. Thirteen studies^13,15,16,18–26,29^ illustrated the potential of PFDs to drive improvements in hospitals and prevent deaths if coroners’ concerns were addressed (Supplement Table S4). For example, lives lost in hospital tragedies, such as the Gosport scandal^31^, could have been prevented if concerns documented in PFDs had been widely disseminated to ensure that actions were taken nationally to safeguard patients.

PFDs have influenced organisations to act, as reported in several studies^18,24,25,28,29^. For example, concerns expressed in PFDs led the Medicines and Healthcare products Regulatory Agency (MHRA) to issue a ‘Drug Safety Update’ Bulletin and give wider publicity to fatal adverse drug reactions^24^. However, research to date has focused on analysing coroners’ concerns in PFDs rather than the actions reported or proposed in responses to PFDs by recipients. Thus, the full potential of PFDs to detect unsafe care and promote change in hospitals is currently underexplored.

### Limitations of PFDs

Several problems in writing PFDs, responding to them, and the ways in which they are written and collected have been highlighted, including inconsistent reporting, poor response rates, and difficulties in accessing and analysing them. Coroners’ practices vary and are not monitored or audited. Three studies^13,23,24^ showed significant variability in the rate at which individual coroners produce PFDs, and six studies^13,15,20,22,23,29^ have reported geographical variation (Supplement Table S4).

The criteria that lead a coroner to write a PFD after an unnatural death, and how coroners decide what information will be included or excluded, are unclear. Two studies^18,22^ showed considerable differences in the quality, detail, length, and structure of reports. Missing information and errors were frequently identified, including failure to disclose the date of birth or age at death, the names or classes of medicines implicated^15,16,20,22,29^, misspelled names, and incorrect dates^13,22^, all of which impair the usefulness of PFDs. In a survey of 32 coroners in 2010-2011, in which they were asked to state and describe their purpose, only six mentioned the importance of preventing deaths as part of their purposes, and only four specifically mentioned PFDs as a tool for doing so^32^. Nevertheless, the attitudes of coroners towards PFDs have improved in the last decade. In the 2020 Coroner Attitude Survey, 96% of coroners responding felt that one of the most important functions of coroners was to ‘prevent future deaths’^33^. More than half of coroners (55%) also felt that PFDs were effective in preventing future deaths^33^.

Given that 16% of all deaths (n=104,940) were considered preventable in Great Britain in 2020^4^ and that only 0.1% resulted in a PFD report^14^, PFDs cannot represent all the actions required to prevent deaths. Cooper et al. highlighted that PFDs report the most severe cases of harm and therefore may not offer generalisable lessons^27^. There is also the problem of timeliness, limiting how swiftly actions can be implemented, owing to the constraints of the coronial system; Ferner and colleagues^25^ found a median delay of 240 days from the date of death to the issue of a PFD. Despite the lag, the concerns in these PFDs still warrant wide public dissemination, to ensure that actions are accountably implemented.

PFDs are unlikely to be highly effective if they are not widely circulated. Responses to PFDs showcase actions taken or proposed that could be implemented nationally or globally to prevent future deaths. However, three studies^24,25,29^ showed that PFDs, which would have benefitted from national dissemination, were most often sent only to local organisations.

Despite the statutory requirement for recipients to respond to PFDs within 56 days^10^, studies reported varyingly low timely response rates between 29 and 37%^15,16,29^. There are no sanctions, penalties, or public accountability for failure to respond to PFDs^23^ and whether actions reported in responses are implemented is unknown^13,15,16,20–22,29^. There is also no mechanism for retaining knowledge and lessons from implementing actions taken after PFDs have been issued. This suggests that there is a need for a two-stage response system where the actions are proposed first in detail then a second response is required sharing how those actions were taken and their results. However, most PFD research has focused on analysing concerns in PFDs rather than the information in responses. Therefore, future research should investigate actions reported or proposed in responses to PFDs.

The impact of PFDs is yet to be fully elucidated, as studies of their effectiveness in improving clinical practice and policy have not been conducted. Several authors^15,16,18,22,24,26,29^ have reported that coroners raised repeated concerns, suggesting that PFDs did not result in the implementation of remedial actions. For example, a case report of two deaths from alcohol-based hand sanitisers illustrates that if lessons had been disseminated nationally and actions had been implemented in all hospitals, such as the use of lockable dispensers, after the first death in 2013, a subsequent death, in 2015, might have been prevented^21^. In some cases, the issues that led to death (e.g. purchasing drugs from the dark web or online pharmacies) are complex^30^ and thus require multifaceted solutions. Before PFDs can be fully incorporated as a patient safety metric for hospitals, future research should assess the effectiveness of PFDs to improve practice and safety and reduce the occurrence of preventable deaths.

The current mechanism of disseminating PFDs on the Courts and Tribunals website is inadequate. To access and analyse information in PFDs requires many hours (and years) of manual labour. Four studies^13,15,16,24^ showed that PFDs were poorly or incorrectly categorised on the Courts and Tribunals website, with 73% of PFDs (=2939) not assigned to any category^13^, creating ambiguities and impairing the ability to compare PFDs with national data (e.g. from the Office of National Statistics) and international data, which use internationally recognised classification systems to report deaths (e.g. the WHO International Classification of Diseases 11th Revision (ICD-11)). After more than five years of using and scraping the Courts and Tribunals website, several issues have emerged, including significant missing information, misspellings, and formatting inconsistencies^13^. Our web scraper and the Preventable Deaths Tracker (https://preventabledeathstracker.net/), which collates PFDs and receives more than 2000 users each month^34^, are helping to streamline analysis and remove some of these problems, to make PFDs more accessible. However, the Chief Coroner’s Office should continue to introduce technologies and systems to improve the dissemination and use of PFDs.

## Discussion

In this narrative review, we identified 17 studies^13,15–30^ that analysed PFDs in England and Wales. PFDs documenting deaths related to medicines and drugs were explored most (47%; n=8), followed by deaths involving healthcare (24%; n=4) and chemical products (12%; n=2). Currently, the strengths of PFDs lie in the great depth and high quality of information PFDs provide on individual deaths and the clear potential of PFDs to prevent future deaths if well implemented. PFDs, however, are currently limited by inconsistent quality and rates of reporting, poor response rates, and challenges in accessing and analysing them.

The Australian and New Zealand National Coronial Information System (NCIS), established in Australia and New Zealand in 2001 and 2007 respectively, has contributed cases to over 200 publications cited in PubMed, nearly 70% of which involved medicines^35^. This displays the potential for PFDs in the UK and should encourage the adoption of similar systems in other countries.

### Strengths and limitations

Our review is limited by the included research. Only two databases, PubMed and medRxiv, were used for screening, although forward and backward screening was performed to improve our search strategy. Included studies explored many important types of PFDs, such as those involving medicines and COVID-19. However, many causes and settings of preventable deaths, such as hospitals, remain unexplored. As a result, our findings reflect only the PFDs so far investigated, and it is unknown where these strengths and limitations extend to all PFDs.

Furthermore, while poor rates of response have been identified, research into responses and the actions they report or propose to PFDs is limited. It is therefore difficult to assess the quality of responses to PFDs and the corresponding effectiveness of PFDs. The current barriers and facilitators to writing and responding to PFDs remain under-researched, preventing the development of potential strategies to improve the PFD system.

### Implications

PFDs could play a crucial role in measuring and improving patient safety and in supporting the goals of the WHO’s Global Patient Safety Action Plan to minimise harms in healthcare. The information in PFDs is the highest quality of evidence available for preventing such deaths, as other study designs, such as clinical trials, are neither feasible nor ethical to perform in such circumstances. Despite the limited quantity of PFDs (0.1% of all deaths)^14^, they offer unparalleled insights into deaths due to patient safety errors in hospitals, by providing details not offered by other safety metrics, such as Serious Incident reports, Never Events, and events recorded on the NRLS. PFDs offer important opportunities for shared information to improve patient safety and could ultimately help prevent future deaths.

Despite the clear role PFDs could play in informing practice, immediate actions are needed to facilitate writing them, to encourage responses to them, and to enhance the ways in which they are reported and implemented.

National guidelines for PFDs should be introduced to address variability in reports and the use of sanctions, penalties, and/or public accountability should be considered to increase response rates. Given that only one PFD was written for every 328 preventable deaths estimated in 2020 (0.29%; 302 PFDs vs. 104,930 preventable deaths^4,11^), the introduction of an inclusive system that requires coroners to report, after every inquest, the actions taken or proposed, would increase the learning from preventable deaths.

In the meantime, technology should be embedded into the PFD system to improve data quality, collection, automation, and timeliness of PFDs. A regular bulletin and analysis of PFDs should be sent to hospitals and general practitioners to increase their use and lessons learnt. Investment in research is needed to examine the facilitators and barriers to writing and responding to PFDs, the effectiveness of PFDs, actions reported or proposed in responses to PFDs and geographical variations. Using PFDs, many important and wide-reaching types of death have been examined, such as those involving medicines and chemical products, have been examined. However, analysis of PFDs and responses detailing deaths in hospitals is an important evidence gap that requires further research.

## Conclusions

Preventable deaths are a measure of avoidable harms due to unsafe care. However, current patient safety metrics fail to facilitate and the implantation of actions to prevent future deaths, particularly in hospitals. Coroners write PFDs to describe concerns from deaths that should be acted upon to avoid further harms. Academic research has recently begun to unlock a wealth of information on patient safety concerns and to identify some of the ways in which PFDs can contribute. While PFDs may have a limited impact on patient safety in hospitals in their current form, a collective effort involving the Chief Coroner’s Office, coroners, policy makers, bereaved families, healthcare professionals, researchers, the media, and the public could restructure and improve the PFD system to unleash their potential. Improved guidance, enforcement of Regulations 29 of The Coroners (Investigations) Regulations 2013, investment in infrastructure and research, and integration into the NHS Patient Safety Strategy are essential for improving PFDs. PFDs offer a unique opportunity to help fulfil the responsibility of hospitals in preventing deaths and ensuring patient safety.

## Data Availability

All data produced in the present work are contained in the manuscript.

## Contributors

BTB screened the studies, extracted relevant data, conducted all analyses, produced all figures and tables, and wrote the first draft of the manuscript. GCR conceptualised and initiated the review; provided supervisory support and edited the first draft of the manuscript. CH and JKA provided supervisory support and oversight. All study authors read, contributed to, and approved the final manuscript.

