## Supplement for "A systematic narrative review of coroners’ prevention of future deaths reports (PFDs): a potential metric for patient safety in hospitals"

**Table S1**: Search strategy conducted in PubMed on 16 November 2022 to identify literature relating to patient safety metrics and to develop Figure 1. Studies were included if they were published after the metrics date of introduction; in 2003 for the National Reporting and Learning System, 2009 for Never Events, and 2010 for Serious Incident reports.

| **Terms** | **Results** |
| --- | --- |
| ("Never event*"[Title/Abstract]) | 446 |
| ("Serious incident*"[Title/Abstract]) | 259 |
| (National Reporting and Learning System[Title/Abstract]) | 97 |

**Table S2:** Search strategy conducted in PubMed on 19 February 2023 to identify literature relating to Prevention of Future Death reports (PFDs).

| **Terms** | **Results** |
| --- | --- |
| "Regulation 28"[Title/Abstract] | 8 |
| prevent future death*[Title/Abstract] | 119 |
| Prevention of Future Deaths[Title/Abstract] | 4 |
| ("Prevention of Future Death Report*"[Title/Abstract]) | 3 |
| coroner report*[Title/Abstract] | 49 |
| ("Regulation 28"[Title/Abstract]) OR (prevent future death*[Title/Abstract]) OR (Prevention of Future Deaths[Title/Abstract]) OR ("Prevention of Future Death Report*"[Title/Abstract]) OR (coroner report*[Title/Abstract]) | 171 |

**Table S3:** Search strategy conducted in medRxiv on 19 February 2023 to identify literature relating to Prevention of Future Death reports (PFDs).

| **Terms** | **Results** |
| --- | --- |
| preventable deaths | 15,787 |

**Table S4:** Summary of the strengths and limitations of included studies investigating coroners’ Prevention of Future Deaths reports (PFDs) indexed in PubMed and medRxiv from inception until 19 February 2023, ordered alphabetically by category.

| **Study ID** | **Population/types of PFDs** | **Strengths** | **Limitations** |
| --- | --- | --- | --- |
| **Medicine and drug-related** | | | |
| Anis et al. 2022^29^ | 113 CVD-related PFDs involving anticoagulants between Jul 2013 and Dec 2019 | PFDs offer lessons for prescribers and policymakers on the safety and appropriate use of therapies. Acting on PFDs at a national level would help to prevent deaths. | PFDs cannot represent all preventable deaths from CVD involving anticoagulants. Data depends on coroners to write and the Chief Coroner’s Office to publish reports and their responses. Key data, including age and drug type, were missing from many PFDs. It was unclear whether changes are being made to incorporate lessons from PFDs. |
| Aronson et al. 2022^30^ | 17 PFDs where medicines or non-medicinal chemicals were purchased online and contributed to death, dated between Nov 2018 and Dec 2019 | PFDs offer healthcare workers, policy makers, and the public lessons regarding purchasing medicines online. | Under-reporting of deaths involving medications purchased online was likely. Solutions to coroners’ concerns can be challenging to implement. |
| Cox & Ferner 2021^26^ | Two PFDs associated with tramadol | Actions on individual cases reported in PFDs could be used to prevent deaths more broadly if communication and adoption were more widespread. | Coroners send reports locally, although the concerns expressed are of national importance. The timeliness and substance of responses were variable. Responses could fail to provide operational guidance to prevent future deaths. |
| Dernie et al. 2022^16^ | 219 PFDs involving opioids | PFDs provide ‘important real-world insights’. If coroners’ concerns, as expressed in PFDs, were addressed, opioids could be used more safely and appropriately. | Issuing PFDs depends on the working practices of individual coroners; the process is neither audited nor assessed. Missing information, such as age at death or date of birth, was identified. |
| Ferner et al. 2019^25^ | Responses to 69 of 99 PFDs relating to medicines, published between Apr 2015 and Sep 2016 from 106 organisations | Concerns raised by coroners are of ‘national importance’. There is evidence that PFDs have prompted actions that otherwise would not have been completed. Coroners often bring to light systemic failures of general importance. | Important concerns are often shared only locally, preventing widespread learning from nationally relevant lessons. Few responses to PFDs are published, and little information is provided as to why. Auditing of responses and their effectiveness is currently an opaque process and, if undertaken, the results are not published. Therefore, it is not possible to determine whether PFDs have saved lives. Often the strategies to prevent future deaths documented in PFD responses do not seem to be effective and applicable. |
| Ferner et al. 2018^24^ | 99 PFDs in which medicines or the medication process was identified, published between Apr 2015 and Sep 2016 | PFDs provide essential information on pharmacovigilance, including medication errors and adverse reactions. Pharmacovigilance could benefit if health organisations, professional and regulatory bodies, and market authorisation holders were more aware of PFDs. | PFDs rarely recognise new hazards. Recipients of concerns are often limited to local organisations, limiting the dissemination of widely relevant lessons. The use of PFDs is impeded by a failure to classify deaths as related to medicines. The working practices of coroners vary significantly, such as to whom they send reports and their criteria for writing PFDs. Coroners and the pharmacovigilance community do not have fully aligned interests. Coroners cannot explicitly make recommendations as to how concerns should be addressed. Limited publication of responses leads to uncertainty as to whether actions have been taken. |
| France et al. 2022^15^ | 704 PFDs involving medicines, published between Jul 2013 and Feb 2022 | Addressing coroners’ concerns could feasibly improve the safety of medicines. PFDs contain important information and have the potential to contribute to a learning environment in clinical practice that could prevent future deaths. | With the same concerns being highlighted repeatedly and a low response rate, lessons are unlikely to be learned. Information, such as age, types of medicines, and dates of responses, were missing from a large proportion of PFDs. The current PFD system does not take steps to enforce or audit responses to PFDs, despite the legal requirement. Concerns should be shared more broadly to ensure that national lessons are learnt. |
| Thomas & Richards 2021^19^ | One PFD attributed to an NSAID, diclofenac | The importance of considering adverse drug reactions from NSAIDs in children and adolescents, especially those with complex needs, was highlighted. | No response to the PFD from the NHS trust had been published 4 years after issue. Responses to PFDs can be opaque, preventing general application of lessons. |
| **Chemical product-related** | | | |
| Bilip & Richards 2021^28^ | One PFD relating to paraffin-based emollient creams | The PFD was sent to several national organisations, including the DHSC, which responded outlining actions previously taken in response to deaths involving paraffin-based emollients. | Further emollient deaths occurred after this PFD, thus, lessons were not learned and actions are still needed. Of the six organisations to which the PFD was addressed, only three responded by January 2021. |
| Richards 2021^21^ | Two PFDs describing deaths from ingesting alcohol-based hand sanitisers | PFDs provide an ‘opportunity to develop and implement mitigation strategies’ and ‘an opportunity to educate healthcare professionals and the public in harm reduction’. Appropriate action in response to deaths could prevent future incidents. | No process is in place to confirm the implementation of actions discussed in addressees’ responses. Coroners’ concerns were not widely communicated in the NHS despite being addressed to NHS England. |
| **Diagnosis-related** | | | |
| Cooper et al. 2021^27^ | Nine PFDs involving diagnostic errors related to GP services in or alongside emergency departments, published between 2013 and 2018 | PFDs often detail root cause analysis and expert opinion, offering an understanding of factors that may have contributed to deaths. | PFDs cover only the most severe cases that have led to death and therefore may not be generalisable to cases of harm without death. |
| **Healthcare-related** | | | |
| King & Benbow 2022^22^ | 159 PFDs in two categories, hospital-related and community health care and emergency service-related deaths, published before May 2021 | PFDs have the potential to stimulate healthcare changes to improve public health and safety. | Details, such as age, were included in PFDs inconsistently, and errors, such as spelling errors and incorrect dates, were noted. The length and detail to which concerns were explained varied. There was wide geographical variation in the writing of PFDs. No studies have been conducted that can demonstrate the ability of PFDs in promoting changes in the healthcare system. The lack of published responses and repeated concerns suggest ineffective action. |
| Leary et al. 2021^18^ | 710 healthcare-related PFDs, published between 2016 and 2019 | PFDs provide ‘valuable insight’, aggregation and analysis of which could inform policy development. Themes arising from PFDs align somewhat with those from non-fatal incident reporting. | PFDs vary significantly in length, scope, and depth of issues raised. Repeated concerns issued to repeated organisations suggest that the lessons offered by PFDs are not always acted on. |
| Swift et al. 2022 ^20^ | 23 PFDs relating to SARS-CoV-2 published between Jan 2020 and Jun 2021 | Coroners’ concerns should be considered during the UK Government’s inquiry into the handling of the pandemic, so that repeated mistakes can be avoided. | There is no formal system by which PFDs are currently audited or analysed, so coroners’ concerns may go unreported and unrecognised. |
| Van Dellen et al. 2022^17^ | The role of psychiatrists in the coronial process | Psychiatrists can identify those to whom PFDs should be addressed. | Thoughtful responses and submissions by psychiatrists can prevent the production of a PFD. This potentially prevents important lessons from being learned and disseminated more widely. |
| **Mixed categories*** | | | |
| Fox & Jacobson 2021^23^ | 50 ‘recently’ published PFDs as of 10 Jun 2020 for categories including child deaths, alcohol, drugs and medications, and railways | PFDs report concerns that, if remedied, could realistically prevent future deaths. These reports can potentially encourage support from bereaved families and the wider public for the process of an inquest and improvement in health and safety. | The statutory duties of both coroners and addressees are not currently being fulfilled. Coroners had varying practices in how they chose to write a report and how they completed them. Coroners are rarely audited and are unable to compare their approaches with those of others. A minority of coroners associated PFDs with their purpose, and even fewer thought that their reports had an active role in public health. |
| Zhang & Richards 2022^13^ | All available PFDs, published between Jul 2013 and Jun 2022 (n=4001) | PFDs hold a ‘rich source of information that decision-makers should use to improve public health and safety’. The introduction of technology can simplify the process of auditing PFDs and ensure that changes are made to prevent future deaths. | One in three PFDs were without response. The Courts and Tribunals website had significant problems with poor categorisation, missing information, and formatting issues; many PFDs contained misspellings and formatting inconsistencies. Data, including sex and age, were also missing from a large proportion of reports. Poor management and standardisation with the Courts and Tribunals website limit the analysis of PFDs. |

PFD: prevention of future death report; CVD: cardiovascular disease; NHS: National Health Service; MHRA: Medicine and Healthcare products Regulatory Agency; DHSC: Department of Health and Social Care. *Categories are determined by the Chief Coroner’s Office when shared on the Courts and Tribunals Judiciary website
